# Plasma Itaconate elevation following successful cDMARD treatment in early rheumatoid arthritis patients elucidates disease activity associated macrophage activation

**DOI:** 10.1101/19001594

**Authors:** Rónán Daly, Gavin Blackburn, Manikhandan Mudaliar, Karl Burgess, Anne Stirling, Duncan Porter, Iain McInnes, Michael P. Barrett, James Dale

## Abstract

**Objective:** To characterize changes in the plasma metabolic profile in newly diagnosed rheumatoid arthritis (RA) patients upon commencement of conventional disease modifying anti-rheumatic drug (cDMARD) therapy.

**Methods:** Plasma samples collected in an early RA randomized strategy study (NCT00920478) that compared clinical (DAS) disease activity assessment with musculoskeletal ultrasound assessment (MSUS) to drive treatment decisions were subjected to untargeted metabolomic analysis. Metabolic profiles were collected at pre- and 3 months post commencement of non-biologic cDMARD. Metabolites that changed in association with changes in the DAS44 score were identified at the 3 month timepoint.

**Results:** A total of ten metabolites exhibited a clear correlation with reduction in DAS44 score following cDMARD commencement, particularly itaconate, its derived anhydride and a derivative of itaconate coA. Increasing itaconate correlated with improved DAS44 score and decreasing levels of CRP.

**Conclusion:** cDMARD treatment effects invoke consistent changes in plasma detectable metabolites, that in turn implicate clinical disease activity with macrophages. Such changes inform RA pathogenesis and reveal for the first time a link between itaconate production and resolution of an inflammatory disease in humans. Quantitative metabolic biomarker based tests of clinical change in state are feasible and should be developed around the itaconate pathway.

**Key Messages:** *What is already known about this subject?:* Rheumatoid arthritis is associated with perturbations in metabolic activity, which have also been associated with response to certain treatments. In vitro work on immunometabolism has recently revealed itaconate as a key metabolite controlling macrophage activation.

*What does this study add?:* In newly diagnosed RA, commencement of csDMARD therapy is associated with changes in the levels of ten metabolites (especially itaconate and its derivatives) that correlate to a corresponding fall in disease activity Pathway analyses suggest these metabolites are associated with macrophage activation.

*How might this impact on clinical practice?:* Changes in metabolite levels in response to treatment provide additional new insights into RA pathogenesis that suggest a focus on macrophage activation state. The association of increased itaconate with decreased inflammation point to possible routes of intervention in RA.

---

Rheumatoid arthritis (RA) is a chronic, destructive, immune mediated inflammatory condition that predominantly affects synovial joints. In genetically susceptible individuals, mucosal exposure to external stimuli (e.g. cigarette smoke) triggers persistent systemic autoimmunity, and subsequent inflammatory cell articular recruitment, leading to tissue damage (1). Constitutional features, such as weight loss, malaise or fever are prevalent in RA patients and RA patients exhibit an increased resting metabolic rate (2), which may in part be related to increased immune cell activation and turnover. Many data now suggest that circulating leukocyte subsets exhibit altered phenotypic and functional properties in the context of RA (3). Chronic synovitis is associated with angiogenesis and consequent increased mediator release e.g. prostanoids and chemokines that may be detected in the circulation. Cardiometabolic disease is a common co-morbidity, reflected particularly in dysregulation of lipid metabolism (4) and has been attributed to an interaction between conventional risk factor pathways and systemic pro-inflammatory cytokines (1). Thus, it is possible that changes in disease activity state may be reflected in measurable changes in biochemical activity that is demonstrated through detailed characterization of metabolite profiles.

Metabolomic technologies provide a detailed description of the relative abundance of individual metabolites within a single tissue or biological system (5,6). At an individual level, these metabolomic ‘signatures’ are the final expression of a complex process of gene-environmental interactions, gene and inflammatory cell activation and protein synthesis (5). Analysing metabolomic profiles across a group of individuals can offer insights into disease pathogenesis when common associations with clinical phenotype emerge. The increased ability to measure quantities of a wide range of different metabolites has permitted detailed description of metabolic profiles across a range of complex, polygenic disorders (7). NMR and LC-MS based metabolomics are being increasingly employed to understand the biochemical changes associated with rheumatoid arthritis (RA) (5,8–15). For example, plasma metabolic profiles, obtained using 1H NMR, differentiated patients with different RA disease activity and showed treatment with TNF-alpha inhibitors modified the baseline metabolic profiles associated with active RA to resemble those of patients in remission (16). Further, Serum metabolite profiles obtained using 1H NMR at baseline and at 24 weeks after treatment also distinguished responders from non-responders to methotrexate treatment (17).

Herein we used an LC-MS platform to characterise changes in the plasma metabolomic profile in newly diagnosed RA patients commencing first line non-biologic conventional disease modifying anti-rheumatic (cDMARD) therapy. Through an untargeted approach we aimed to determine whether the levels of individual metabolites correlated to disease activity following initiation of treatment and whether changes in disease activity were also reflected in changes in the level of certain metabolites.

## Patients and Methods

### Study Population

This study was conducted using clinical data and samples from 79 patients recruited to the Targeting Synovitis in Early Rheumatoid Arthritis (TaSER) study, a randomised clinical trial that compared the effectiveness of using either clinical (DAS28) or musculoskeletal ultrasound (MSUS) assessment of disease activity to drive an intensive early treatment strategy (18,19). Briefly, at recruitment, all patients had active RA (DAS44>2.4) and both groups followed the same step-up sequence of DMARD escalation. In the DAS28 group, treatment was escalated until low disease activity was attained (DAS28<3.2) and in the MSUS group treatment was escalated until 1 or no joints of a limited 14 MSUS joint set exhibited any power Doppler (PD) signal. At the start of treatment patients were treated with methotrexate, or sulphasalazine if methotrexate was contraindicated, and combinations of intra-articular and intra-muscular corticosteroids. Disease activity assessments, using the 44-joint disease activity score (DAS44), were conducted at baseline and every 3 months by a metrologist (AS) who was blinded to group allocation and treatment. The earliest that study group allocation could influence ongoing treatment was after 3 months of follow-up. The study protocol was approved by the West of Scotland Research Ethics Service and was registered with ClinicalTrials.Gov (NCT00920478). All patients provided written consent to participate and for their disease activity results and tissue samples to be used for research purposes. All study activities were conducted in accordance with the Declaration of Helsinki.

### Sample Collection and Preparation

All patients donated additional blood samples for research purposes at baseline (group A), 3 months (group B) and 18 months of follow-up (group F) using a standard operating procedure for sample harvest and processing. Blood was collected into lithium heparin vacutainers and stored on ice. Samples were centrifuged at 4°C (1100g, fixed angle rotor) within 4 hours of venipuncture and 500ul aliquots of plasma were stored at −80°C until required for analysis.

### Metabolomics

Samples were analysed by hydrophilic interaction liquid chromatography (HILIC) -mass spectrometry (LC–MS) (UltiMate 3000 RSLC (Thermo Fisher, San Jose, California, USA) using a 150 × 4.6 mm ZIC-pHILIC column (Merck SeQuant, Umea, Sweden) running at 300 ll/min and Orbitrap Exactive (Thermo Fisher) detection. Mass spectrometer parameters were: 50,000 resolving power in positive/negative switching mode. Electrospray ionisation (ESI) voltage was 4.5 kV in positive and 3 kV in negative modes. Buffers consisted of A: 20 mM ammonium carbonate in H2O and B: Merck SeQuant: acetonitrile. The gradient ran from 20 % A: 80 % B to 80 % A: 20 % B in 900 seconds, followed by a wash at 95 % A: 5 % B for 180 seconds, and equilibration at 20 % A: 80 % B for 300 seconds.

The LC-MS data was processed using a combination of open source tools run though R. Vendor-specific raw LC-MS files were converted into the mzXML open format using MSConvert from the proteowizard pipeline (20). During conversion the m/z data was centroided. Chromatographic peaks were extracted from the mzXML files using the centwave detection algorithm from XCMS and converted to PeakML files. Subsequently, PeakML files representing replicates were aligned and combined using mzMatch.R (21) after filtering out all peaks that were not reproducibly detected within groups. The combined PeakML files were subjected to additional intensity filtering, noise filtering and gap-filling to produce a set of reproducible peaks. These peaks were then corrected for instrument drift over time using an in-house Gaussian process regression algorithm modelled on the pooled samples. Peaks were manually checked for consistency and integrated using QuanBrowser (Thermo Fisher) where appropriate. Identifications were based on the Metabolomics Standards Initiative proposed minimum reporting standards.

### Statistical Analysis

Demographic and disease activity outcome data was collected from the TaSER study records. Tests of significant differences between peak levels were calculated using t-tests, and controlled for by correcting the p-values for multiple testing by calculating q-values. Relationships between disease activity and metabolite levels were modelled using linear regressions and tests of significance were controlled by calculating q-values. Correlation coefficients were calculated using Pearson’s product-moment method. Full metabolome correlation analysis was performed using partial least-squares analysis of the full set of features on DAS44. Statistically highlighted features were manually assessed for peak shape to determine if they correspond to genuine metabolite related signals. Metabolite identification was carried out by first calculating an accurate molecular formula for the m/z value within 3ppm. This formula was then compared to a list of authentic standards and assigned as a match if the retention time and peak shape were comparable. The list of authenticated standards is included as supplementary material (**Supplementary File 1**). If not found in the authentic standards a putative assignment was made based on the retention time of the feature and the chemistry of the LC column using a curated list of 41,623 metabolites contained within the IDEOM database (http://mzmatch.sourceforge.net/ideom.html), as detailed by Creek et. al. (22). The study dataset has been uploaded to the online Metabolights repository (www.ebi.ac.uk/metabolights)

## Results

### Study Population

Table 1 summarises the baseline characteristics of the patients included in this study. At baseline, 75 patients commenced methotrexate and 4 commenced sulphasalazine After 3 months follow-up we detected a significant improvement in disease activity, with a mean reduction in DAS44 from baseline of 2.1 (SD 1.4). Thirty-five patients were exposed to corticosteroid treatment (1 oral, 9 intra-articular only, 19 intra-muscular only, 6 intra-articular and intra-muscular) prior to donating research blood samples.

**Table 1.**
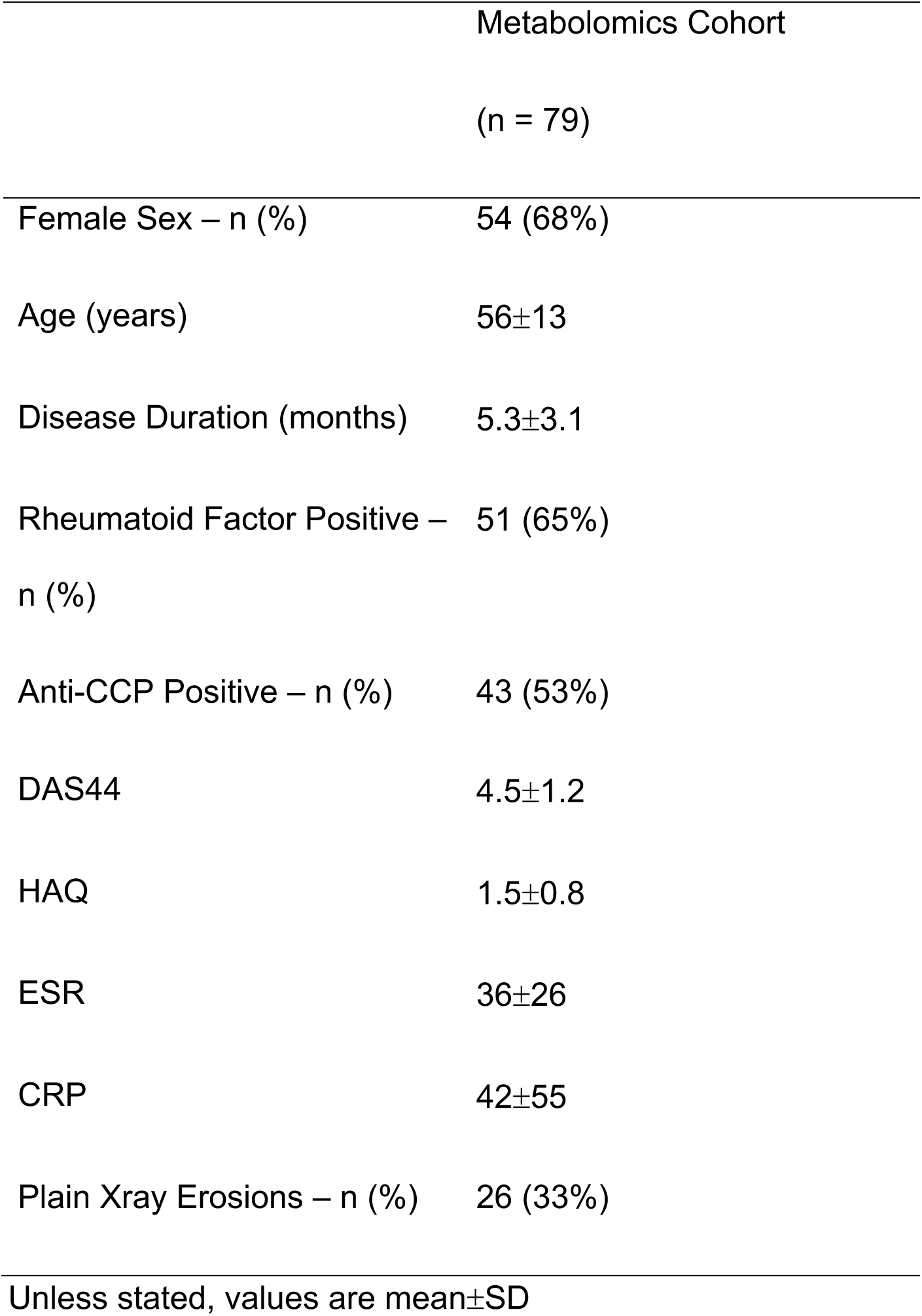
Baseline Characteristics of Study Cohort

### Metabolomic analysis

Plasma from patients was subjected to untargeted metabolomics analysis using LC-MS (23). A PCA plot (**Supplementary File 2**) reveals no appreciable global separation from baseline to 3 months. Nevertheless some individual peak changes were evident and those relating to the biggest differentiators in the PCA loadings were checked against lists of common contaminants (24) and assessed chromatographically as a safeguard against the observed separation being due to a sample handling/processing factor.

### Metabolite Comparisons

Comparisons were performed between individual peaks at baseline and 3 months, to see if there were any significant differences. These comparisons used a basic t-test to calculate a p-value and log fold-change difference. The p-values were used to control the false discovery rate, by calculating q-values. Those differences with a value of q < 0.05 were reported as significant. Out of 3042 peaks in the dataset, 464 were reported as different between baseline and 3 months. These values can be seen in the accompanying spreadsheet (**Supplementary File 3**).

### Relationship between changes in metabolite levels and changes in DAS44

To determine whether changes in disease activity were matched by changes in metabolomic profile, differences in DAS44 and metabolite levels between time points were calculated, for all patients. A linear regression was then performed, regressing DAS44 change on change in peak levels, for the baseline to 3 months. Each set of regressions admitted an effect size and p-value. These p-values were used to control the false discovery rate by calculating q-values.

Between baseline and 3 months, 9 significant effects were found for values of q < 0.05. A volcano plot of all the peaks is shown in Figure 1, with information on the significant peaks in Table 2. Indicative plots of these data and models are shown in Figure 2. E.g. looking at the model for Peak.n.724, the slope of the line is −0.5, which indicates that a doubling/halving of the concentration after treatment, is associated with an extra change in DAS44 downwards/upwards of approximately 0.5. This extra change is on top of the average DAS44 change in the whole population. Once identified as significant effects, these signals were manually assessed to determine peak quality and identity (where possible).

**Table 2.**
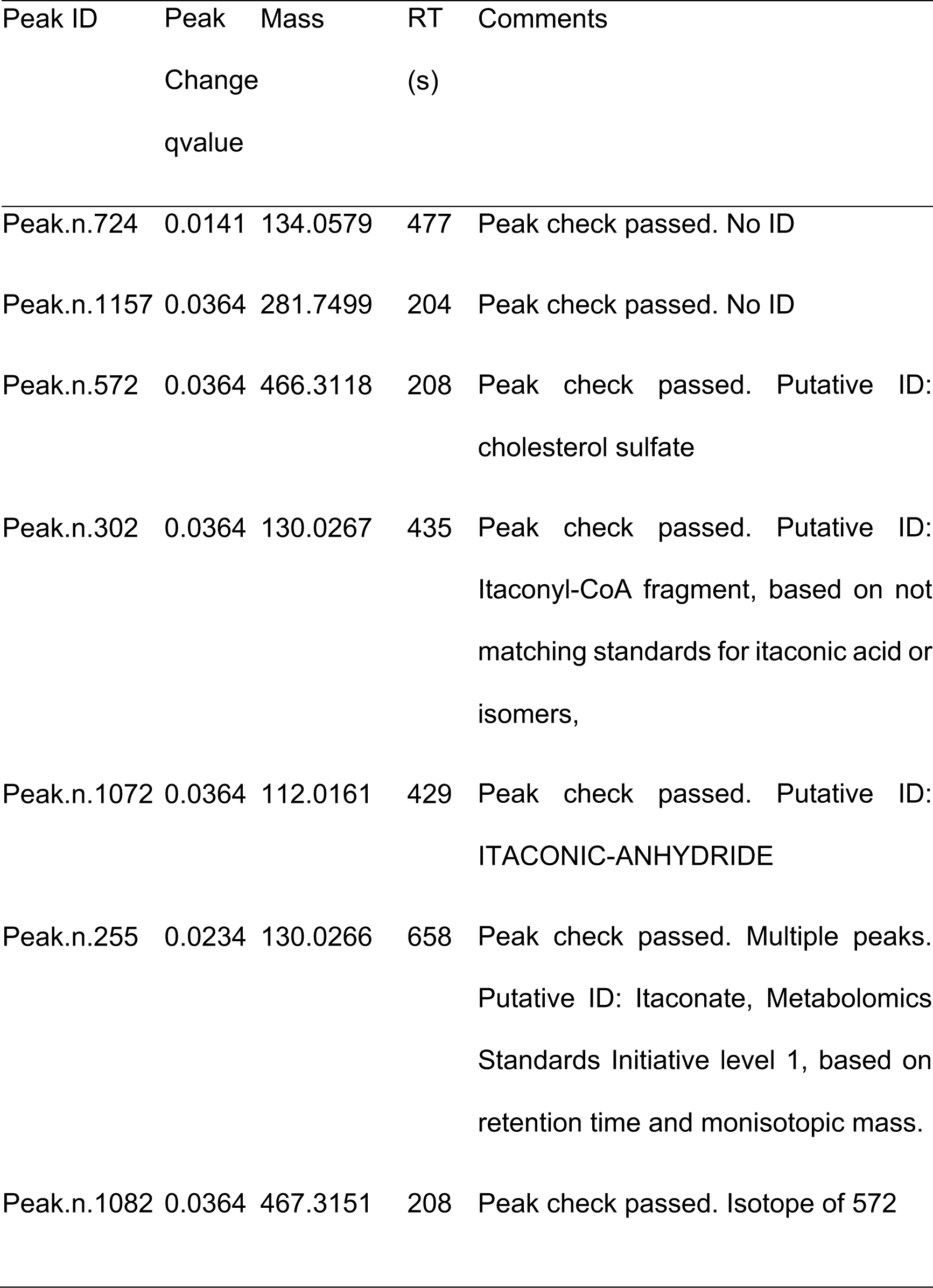
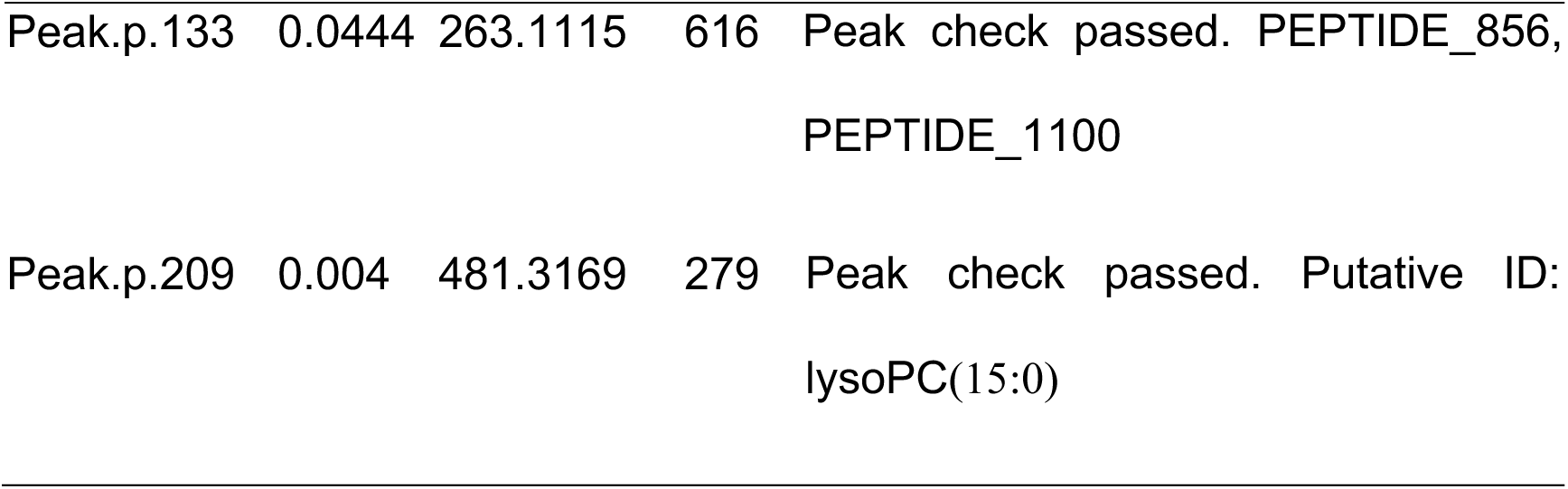
Annotated LC-MS peaks that have been differentially expressed across changing DAS44 scores.

**Figure 1.**
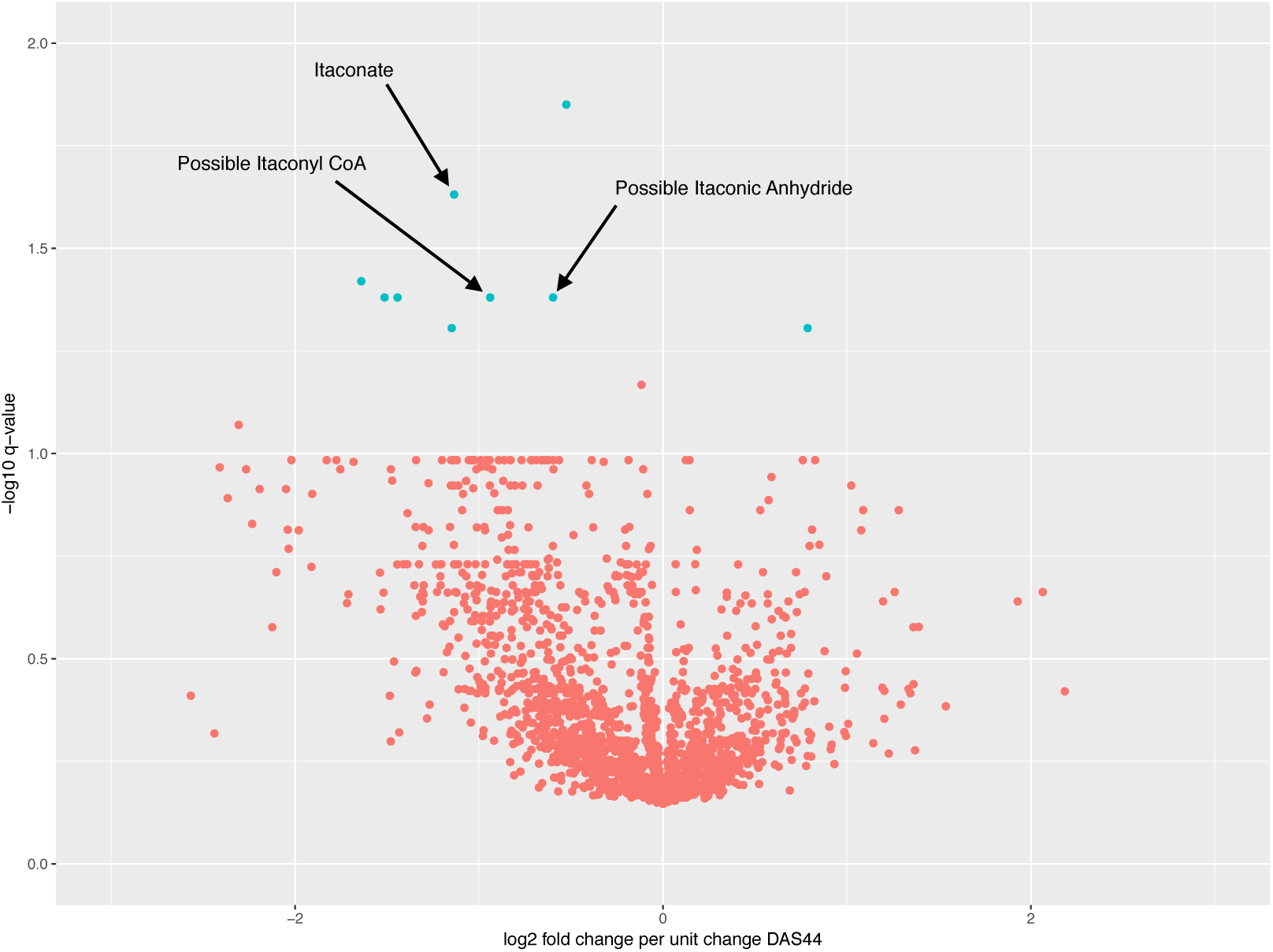
Volcano plot between baseline and three months. Blue points are significant peaks.

**Figure 2.**
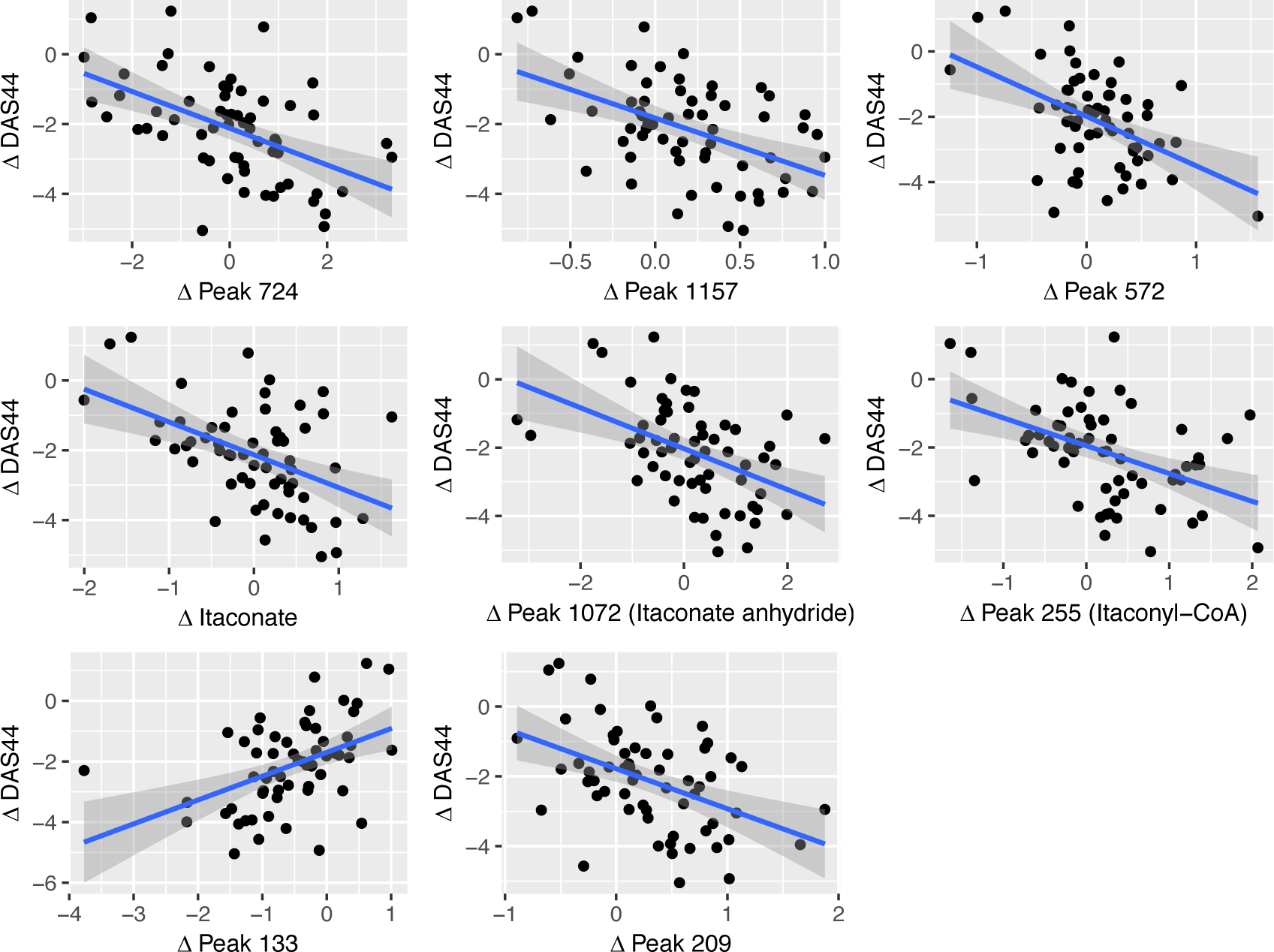
Scatter plots demonstrating change in DAS44 between baseline and 3 months vs change in log_2_ peak intensity of 8 putative metabolites. The Itaconate peak has been identified. Peak 1072 and 302 have been given putative identities of Itaconate anhydride and Itaconyl-CoA respectively.

Foremost among those metabolites associated with the decline in DAS44 score were itaconate (mz = 130.0267), a predicted itaconate anhydride (112.016) and a fragment predicted as originating from itaconyl coA (130.0266) (Figure 3). Among the other metabolites were cholesterol, several peptides and a range of fatty acids.

**Figure 3.**
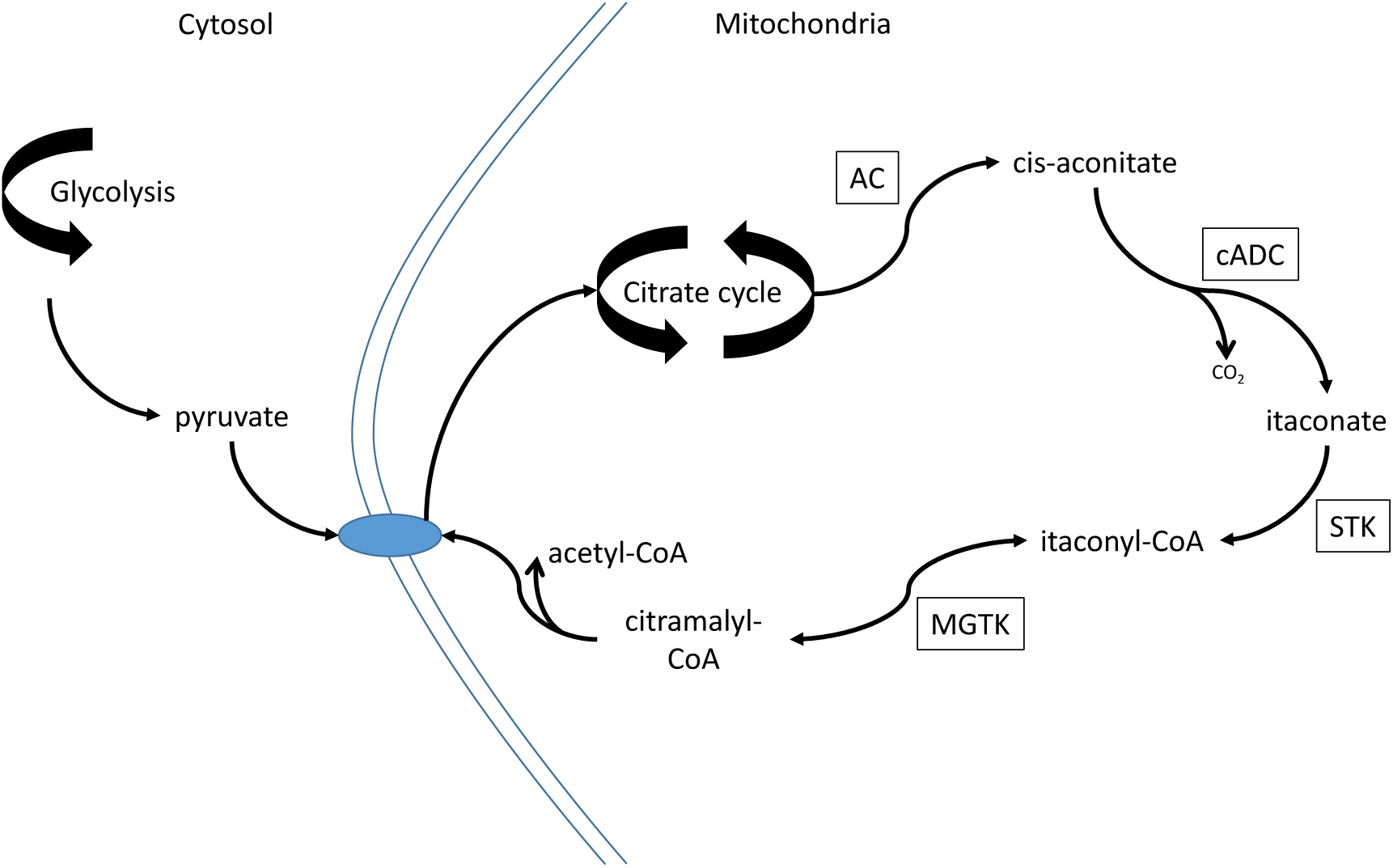
Metabolic pathway showing the production and itaconate via the TCA cycle and metabolism to pyruvate via itaconyl-CoA and citramalyl-CoA. Cis-aconitate is produced from citrate and iso-citrate by aconitase (AC). This is converted to itaconate by cis-aconitate decarboxylase (cADC) which is then converted to itaconyl-CoA via succinate thiokinase (STK). Itaconyl-CoA is converted to citramalyl-CoA by methylglutaconyl-CoA hydratase (MGTK) which is then converted to pyruvate and acetyl CoA (34).

In order to verify the feature-by-feature analysis, a partial least-squares (PLS) analysis was performed on the peak change of the full set of features against change in DAS44. This analysis indicated one component and zero orthogonal components. The Q2Y metric was given as 0.125, with a p-value of 0.01 after 1000 permutations of the samples. The top 9 features from this analysis corresponded exactly to the 9 features given by the feature-by-feature analysis.

These results were then checked against those peaks that had a significant difference between baseline and 3 months, to find those peaks where there was both a significant difference between peak levels in the population, and also where there was a correlation between the change in DAS44 and the difference in peak levels. There were three peaks with this property, Peak.p.133 (annotated as ser-ser-ala or gly-ser-thr), Peak.p.209 (annotated as LPC(15:0)) and Peak.n.1157 (mass of 281.7500, retention of 203 s). n.1157 was not matched to any known metabolite, it presents as a doubly charged [M-2H-]2-peak with a predicted formula of C30H61N9O.

### Itaconate and CRP level have similar predictive power for response

Blood CRP levels are measures of the acute phase response that have rapid change properties that can map with response to treatments. Accordingly it is included as an indirect surrogate of immune cell activation and has been included in a number of composite disease activity measures such as DASCRP-28 and SDAI. In this study, change in CRP levels correlated positively with change in DAS44 score (r=0.41, p=9.4×10^−4^), diminishing as disease activity reduced. Conversely, change in itaconate correlated negatively with change in DAS44 (r=-0.49, p=9.6×10^−5^). Besides being correlated with DAS44, CRP and itaconate are also negatively correlated (r=-0.44, p=4.9×10^−4^). These associations are shown in Figure 4.

**Figure 4.**
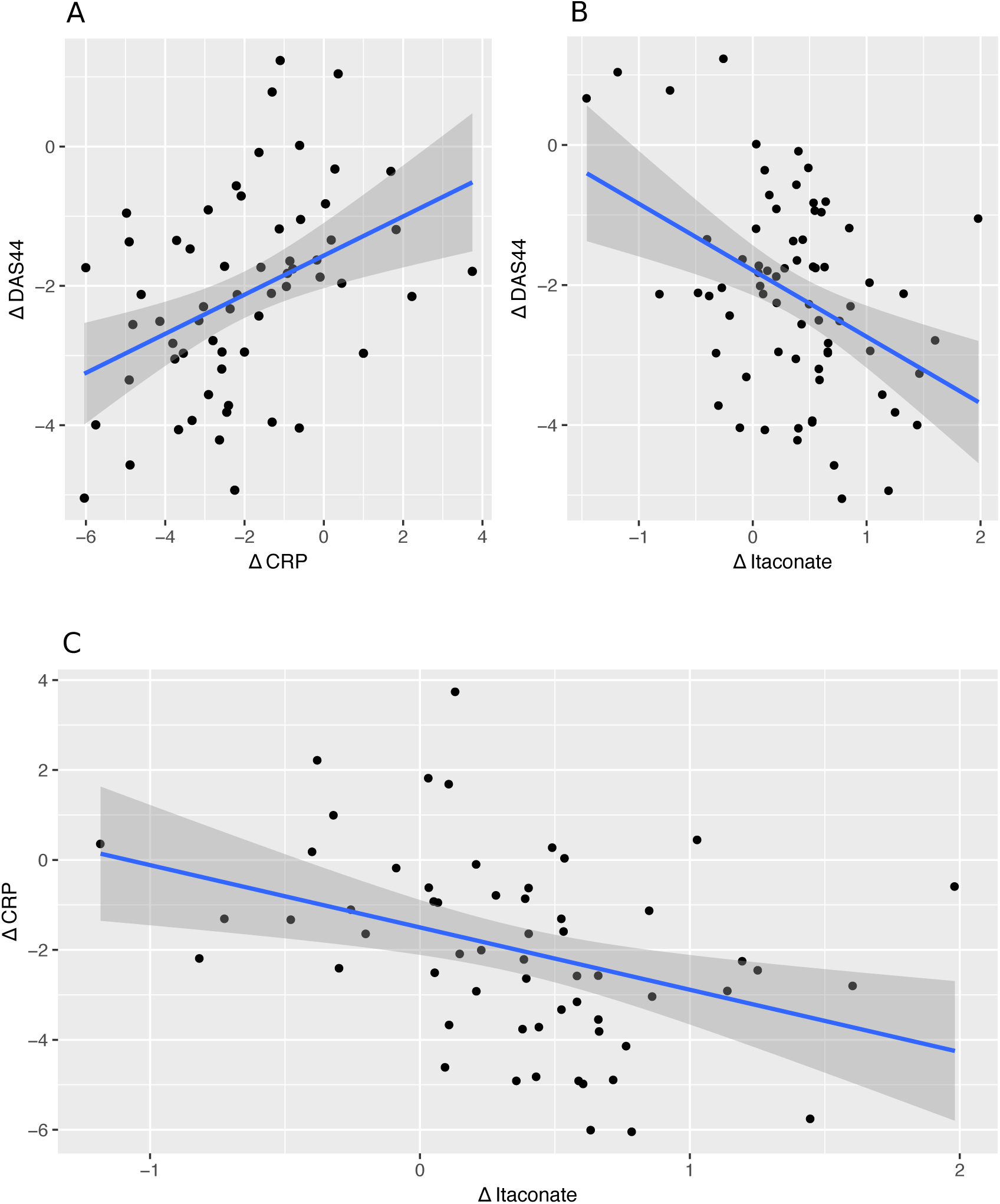
Scatter plots showing correlation between A) change in CRP between baseline and 3 months vs change in DAS44 between baseline and 3 months (r = 0.41); B) change in itaconate between baseline and 3 months vs change in DAS44 between baseline and 3 months (r = −0.49); C) change in itaconate level between baseline and 3 months vs change in CRP between baseline and 3 months (r = −0.44)

## Discussion

Metabolomics is emerging as a tool to identify biomarkers for disease, response to treatment and also indicators of pathogenesis that may offer routes for novel interventions. In the past few years a number of studies using both NMR and mass spectrometry based approaches have been applied to study RA (5–17,25–27). The use of an untargeted LC-MS platform has several benefits over the use of other platforms, such as NMR. Whilst the reproducibility and quantitation provided by NMR allows standardization across laboratories, the technique is hampered by poor sensitivity and the inevitable overlap of strong signals (such as water) with weak ones (including many metabolites of interest). Mass spectrometry is capable of detecting, and identifying, a much broader range of metabolites and combining it with a suitable LC set up optimised for small, polar metabolites will provide a much more complete picture of the metabolome (23). Mass spectrometry also allows for better identification of unknown metabolites; signals that do not match a known metabolite in any of the databases (28). We therefore adopted this approach to seek novel biomarkers of state and response in patients with new onset RA.

Our results demonstrate a clear association between short term changes in DAS44 scores and levels of a panel of 9 metabolites. Principal amongst these is itaconate, its derived anhydride and a putative itaconate coA derivative. Further, the correlation between DAS44 score and itaconate is slightly more robust than that between DAS44 score and CRP, indicating the itaconate might be as good a marker of improved patient status as CRP (both, however, remaining less good than DAS44 score alone).

Itaconate has recently emerged as a primary moiety of interest in the pathogenesis of inflammatory disease and macrophage activation to an M1 / inflammatory phenotype. Initially an ability to inhibit succinate oxidation by succinate dehydrogenase was proposed as the key driver on its immunomodulatory effect (29). Recently however, it has been shown that itaconate impacts directly on the anti-inflammatory transcription factors Nrf2 (30, 31) to underpin its immunomodulatory activity. Itaconate is also the most pronounced marker of inflammatory arthritis in a murine model (32). A recent study in the Tg197 murine model of inflammatory arthritis described itaconate to be a key marker of the disease, and elevated levels were found in afflicted mice that reversed upon TNF blockade with infliximab (32). In other studies, itaconate appears to be involved in regulation of inflammation, its elevation leading, ultimately, to suppression of the inflammatory response (33) and although functional roles in human inflammatory disease have yet to be reported, our studies indicate levels are elevated as inflammation is diminished. Our data reveals increasing itaconate associated with decreasing DAS score after cDMARD (primarily methotrexate) treatment, which is consistent with its anti-inflammatory role. However, the analysis in Tg197 mice indicated that elevated levels of itaconate were found during disease manifestation and these declined upon anti-inflammatory treatment with the anti-TNFα monoclonal antibody infliximab. This observation appears to be contradictory to ours. However, a number of differences between the studies need to be considered. Our work used human plasma while hind limb tissue extracts and synovial fibroblast extracts were found to be the optimal material to find metabolite changes the transgenic mouse model. Our first comparison point to baseline came at three months while the mouse comparator point was at six weeks and our study did not include a cohort of healthy controls to compare itaconate levels at the start of the experiment. Comparing cDMARD based therapy and biological-based therapy may be another confounding factor since different therapies work via different mechanisms to diminish inflammation and therefore may have differing influences on itaconate levels. Notwithstanding, having demonstrated a clear link between itaconate levels and response to treatment in RA, future studies using a wider range of variables will ultimately reconcile our understanding of the role of this pathway in inflammatory disease.

This study cannot distinguish whether itaconate production is a result of improved condition due to treatment or is in itself responsible for that improved condition. Nor can it determine what impact exposure to specific treatments has on itaconate expression. The initial observation requires validation in an independent RA cohort and within cohorts of patients with other inflammatory diseases. However, as the first indication that there is a clinically evident association between itaconate levels and disease activity levels, the need for additional work to understand if stimulating itaconate production pharmacologically offers a route to intervention in RA is of major importance. The findings suggest that further study of the itaconate pathway and macrophage activity may reveal additional important insights into immune function regulation and RA pathogenesis and may also reveal new, clinically relevant, markers of disease activity and treatment response. Finally, this study provides proof of concept that additional insights to disease pathogenesis can be identified through analysis techniques that combine highly detailed descriptions of metabolite expression with clinical data.

## Data Availability

Please contact the corresponding author in regards to accessing the experimental data.

## Notes

Funders: The TaSER study was jointly funded by a Clinical Academic Fellowship from the Chief Scientist’s Office, Scottish Executive and an Investigator Initiated project grant from Pfizer UK. The metabolomics study was funded by an additional Investigator Initiated project grant from Pfizer UK. RD was funded by Wellcome (105614/Z/14/Z). MPB is part of the Wellcome Centre for Molecular Parasitology funded by a core grant from Wellcome (104111/Z/14/Z).

Competing Interests: RD, GB, KB, DP and MB have nothing to disclose. MM reports other support from CEREVANCE, LTD, outside the submitted work. AS reports grants from Pfizer UK, during the conduct of this survey. IM reports grants from Pfizer, grants from Chief Scientist Office, grants from ARUK, during the conduct of the study; grants and personal fees from BMS, personal fees from Abbvie, grants and personal fees from UCB, grants and personal fees from Pfizer, grants and personal fees from Janssen, personal fees from Lilly, outside the submitted work. JD reports grants from Chief Scientist’s Office, Scottish Government and from Pfizer UK during the conduct of the study; non-financial support from Abbvie UK, outside the submitted work.

### Competing Interest Statement

RD, GB, KB, DP and MB have nothing to disclose. MM reports other support from CEREVANCE, LTD, outside the submitted work. AS reports grants from Pfizer UK, during the conduct of this survey. IM reports grants from Pfizer, grants from Chief Scientist Office, grants from ARUK, during the conduct of the study; grants and personal fees from BMS, personal fees from Abbvie, grants and personal fees from UCB, grants and personal fees from Pfizer, grants and personal fees from Janssen, personal fees from Lilly, outside the submitted work. JD reports grants from Chief Scientist’s Office, Scottish Government and from Pfizer UK during the conduct of the study; non-financial support from Abbvie UK, outside the submitted work.

### Clinical Trial

NCT00920478

### Funding Statement

The TaSER study was jointly funded by a Clinical Academic Fellowship from the Chief Scientist?s Office, Scottish Executive and an Investigator Initiated project grant from Pfizer UK. The metabolomics study was funded by an additional Investigator Initiated project grant from Pfizer UK. RD was funded by Wellcome (105614/Z/14/Z). MPB is part of the Wellcome Centre for Molecular Parasitology funded by a core grant from Wellcome (104111/Z/14/Z).

### Author Declarations

All relevant ethical guidelines have been followed and any necessary IRB and/or ethics committee approvals have been obtained.

Any clinical trials involved have been registered with an ICMJE-approved registry such as ClinicalTrials.gov and the trial ID is included in the manuscript.

